# Calmodulin variants in schizophrenia patients display gain-of-function or loss-of-function effects

**DOI:** 10.1101/2024.05.22.24307674

**Authors:** Helene Halkjær Jensen, Malene Brohus, John W. Hussey, Ana-Octavia Busuioc, Emil Drivsholm Iversen, Faezeh Darki, Gabriela Dobromirova Nikolova, Amalie Elton Baisgaard, Palle Duun Rohde, Ida Elisabeth Gad Holm, Andrew McQuillin, Torben Moos, Ivy E. Dick, Michael Toft Overgaard, Mette Nyegaard

**Author notes:** Authors contributed equally (Helene Halkjær Jensen, Malene Brohus). Authors contributed equally (Michael Toft Overgaard, Mette Nyegaard). Correspondence: Helene Halkjær Jensen, Department of Chemistry and Bioscience, Aalborg University, Fredrik Bajers Vej 7H, 9220 Aalborg Ø, Denmark; Mette Nyegaard, Department of Health Science and Technology, Aalborg University, Selma Lagerløfs vej 249, 9220 Aalborg Ø, Denmark.

## Abstract

Calmodulin acts as a vital calcium sensor in cells, crucial for relaying calcium signals to different protein partners. While rare missense variants in calmodulin are linked to cardiac arrhythmia, particularly long QT syndrome (LQTS), their role in schizophrenia remains unexplored. We investigated missense variants in the calmodulin-encoding genes *CALM1-3* in a large-scale sequencing effort involving 24,248 schizophrenia patients and 97,322 controls. Seven carriers were found among cases and twenty among controls. Notably, all schizophrenia variants affected the C-terminal lobe of the protein, compared to only five in controls, linking calmodulin C-lobe missense variants and schizophrenia risk (odds ratio 5.62, *P*=0.043). Functional analyses revealed two classes of calmodulin variants in schizophrenia: 1) loss-of-function variants that reduce calcium affinity and impair the interaction with voltage-gated calcium channel 1.2 (Ca_V_1.2), akin to LQTS variants but with smaller effect size, and 2) gain-of-function variants that unexpectedly enhance calcium affinity with no impact on Ca_V_1.2 gating. This study for the first time statistically and functionally links calmodulin missense variants to a neurological disorder, expanding the phenotypic spectrum of calmodulinopathies to include schizophrenia.

**Significance:** Neurons use calmodulin to monitor calcium signals and modulate hundreds of target proteins, thereby regulating key processes such as neuronal firing and memory and learning. Here, we link genetic variants in calmodulin to schizophrenia risk. Moreover, these genetic variants have divergent consequences for calmodulin protein function. Our results expand the current understanding of calmodulin variants, which are primarily reported in cardiac arrhythmia patients, and generally have a strong loss-of-function effect on the protein. We here provide the first identification and characterization of calmodulin variants in non-cardiac patients. This broadens our view of the physiological and functional consequences of human calmodulin variants and presents novel mechanistic entries to understanding the molecular mechanisms of schizophrenia.

## Introduction

Calmodulin is a small calcium binding protein expressed in all cells. When calcium ions enter the cytosol as a cellular signal, calmodulin binds the ions and changes conformation, enabling calmodulin to bind and regulate hundreds of target proteins (1). Calmodulin thus functions as a universal calcium sensor. The protein has four calcium binding sites organized in two lobes; the N-terminal lobe (N-lobe) and the C-terminal lobe (C-lobe). The N-lobe has lower affinity and faster kinetics than the C-lobe, enabling calmodulin to capture calcium changes over a broad temporal and concentration range. Remarkably, calmodulin is exceptionally well-conserved, and the protein sequence is identical in all vertebrates (2). In humans, calmodulin is encoded by three independent genes, *CALM1-3*. These genes have very different intronic and untranslated sequences, but the coding parts produce identical calmodulin proteins in all three genes (3).

In 2012, we were the first to identify missense variants in calmodulin and causally link these genetic variants to cardiac arrhythmia in children (4). This link has since been confirmed in several other studies and explained molecularly (1, 5). Today, patients with disease-causing calmodulin variants are described in the International Calmodulinopathy Registry (ICalmR) (6, 7). In 2023, the ICalmR covered 140 patients worldwide, thus demonstrating that calmodulin variants are ultra-rare. The majority of calmodulinopathy patients suffer from long QT syndrome (LQTS). In these patients, most calmodulin variants affect amino acid residues directly involved in calcium coordination, and almost all variants impair calcium binding in the calmodulin C-lobe (1, 8). Moreover, calmodulin variants in LQTS patients affect calmodulin’s ability to regulate the voltage-gated calcium channel 1.2 (Ca_V_1.2) (9–12). Recently, the phenotypic spectrum of calmodulinopathies has expanded to include structural heart disease as well as developmental delay, but systematic studies on the effect of these variants on Ca^2+^ sensing and Ca_V_1.2 function are lacking (7).

Ca_V_1.2, encoded by the *CACNA1C* gene, is a plasma membrane calcium channel expressed in a number of tissues, including cardiac myocytes and neurons (5). When active, Ca_V_1.2 forms a multi-domain complex consisting of the *CACNA1C*-encoded pore-forming α_1_ domain, an intracellular β subunit, and an extracellular α_2_δ subunit. Calmodulin interacts with an IQ motif in the intracellular C-terminal of the α_1_ subunit (13). When Ca_V_1.2 opens in response to an action potential, calcium ions enter the cell, and calmodulin registers the increased calcium concentration. Calcium binding to calmodulin induces a conformational change that propagates to the Ca_V_1.2 channel and signals channel closure; a process called calcium-dependent inactivation (CDI) (14). Calmodulin variants in LQTS patients reduce or completely abolish CDI (15). In this way, a loss-of-function variant in calmodulin results in a gain-of-function effect on Ca_V_1.2 calcium flux. Thus, calmodulin can be regarded a key subunit in the Ca_V_1.2 channel complex.

Interestingly, genetic variants in *CACNA1C* are associated with both cardiac and neurological disease. In the heart, variants in *CACNA1C* cause several cardiac arrhythmias, including short QT syndrome, Brugada Syndrome, and long QT syndrome type 8 (LQT8) (5). In Timothy Syndrome patients, variants in *CACNA1C* cause LQT8 in combination with developmental defects, syndactyly, and autism spectrum disorder (16, 17). *CACNA1C* variants have also been found in patients with neurological disorders alone, including developmental delay, epilepsy, and intellectual disability (5, 18). We established the first association between *CACNA1C* and schizophrenia (SCZ) in 2010 (19). Since then, variants in *CACNA1C* have frequently been identified as genetic risk factors in genome-wide association studies of SCZ (5, 19–21).

Ca_V_1.2 regulation by calmodulin is equally critical in the brain and in the heart (22), and an increasing number of patients in the ICalmR have been reported with neurodevelopmental disorders (7). Since *CACNA1C* is a SCZ gene, and since calmodulin is a key regulator of Ca_V_1.2, we hypothesized that calmodulin variants are associated with SCZ. To address this hypothesis, we accessed data from a recently published exome sequencing study of a large population of SCZ patients and control individuals (23). In this population, we found seven calmodulin missense variants in SCZ patients and 20 missense variants in control individuals. Remarkably, all SCZ variants were exclusively observed in the C-lobe. Further, we found a significantly higher proportion of SCZ variants affecting calcium affinity compared to control variants. Two of the SCZ variants followed a mechanistic paradigm mirroring LQTS with impaired calcium and Ca_V_1.2 IQ-domain binding, but with smaller effect size than that of LQTS variants. On the other hand, three SCZ variants displayed a new mechanistic pattern with increased calcium affinity that may act through alternative mechanisms to Ca_V_1.2 dysregulation. Together, this study identifies a new phenotypic manifestation of calmodulin variants and proposes that calmodulin variants in part act through other pathways in the brain than in the heart.

## Materials and Methods

### Identification of CALM missense variants in SCHEMA

The Schizophrenia Exome Sequencing Meta-analysis (SCHEMA) has collected full exome or whole genome sequences from 11 international studies of patients diagnosed with schizophrenia or schizoaffective disorder (23). Control subjects were recruited from the same studies as well as from non-psychiatric and non-neurological collections in the Genome Aggregation Database (gnomAD). Control individuals in SCHEMA are thus a combination of individuals with no known psychiatric diagnosis and individuals selected from other population registries. Summary-level data from SCHEMA is available through https://schema.broadinstitute.org. Here, we searched for the genes *CALM1*, *CALM2*, and *CALM3* (last accession on 22^nd^ of September 2023). In the resulting list of genetic variants, we filtered for “missense only”. For all missense variants, we extracted information of their position in the gene, the character of the variant, MPC and CADD score (available when clicking on each variant), and whether they were observed in the patient population or in the control population. Other participant information is not available through the browser, and the data is effectively anonymized.

### GTEx accession

To assess *CALM1-3* gene expression across brain regions, we accessed the Genotype-Tissue Expression (GTEx) portal V8 (24). To extract short read RNA sequencing data, we used the multi gene query through https://www.gtexportal.org/home/multiGeneQueryPage, last on 30^th^ October 2023. Here, we searched for the genes *CALM1, CALM2, CALM3, CACNA1C, CACNA1G, CLDN4, CUL1, GRIA3, GRIN2A, HERC1, RB1CC1, SETD1A, SP4, TRIO, UBC,* and *XPO7.* In the search, we included all available brain regions (amygdala, anterior cingulate cortex (BA24), caudate (basal ganglia), cerebellar hemisphere, cerebellum, cortex, frontal cortex (BA9), hippocampus, hypothalamus, nucleus accumbens (basal ganglia), putamen (basal ganglia), spinal cord (cervical c-1), and substantia nigra), lung (an epithelial tissue), heart left ventricle, and skeletal muscle. We extracted the median transcript-per-million (TPM) for all genes in all tissues.

### Human brain tissue

Brain samples containing hippocampal tissue were obtained from autopsy following written consent by the next of kin, according to the Danish legislation. We used tissue from two adults (50-65y, both male) with no reported neurological disorders. The tissue was fixed in 4% paraformaldehyde for four weeks and embedded in paraffin.

### RNA in situ hybridisation

The brain tissue was cut on a microtome at 3 µm thickness and de-paraffinated in Neo-Clear (109843, Merck). Deparaffination and *in situ* hybridization was performed using the RNAscope® 2.5 HD Detection Reagent RED kit (322360, Advanced Cell Diagnostics), following the manufacturer’s recommendations. Hybridization was performed for 2h at 40°C in a HybEZ™ II Hybridization Oven (321720, Advanced Cell Diagnostics) with probes to detect *CALM1* (461121), *CALM2* (499931), *CALM3* (1128281), *UBC* (positive control, 312028), and *DapB* (negative control, 310043) (all Advanced Cell Diagnostics). Following development of the Fast Red chromogenic signal, slides were counterstained with Mayer’s hematoxylin (MHS16, Merck) for 2 min. Slides were airdried for 15 min at 60°C before mounting in VectaMount® Permanent Mounting Medium (H-5000, Vector Laboratories). Imaging was performed on a Hamamatsu NanoZoomer-S360 Visiopharm slide scanner with identical settings for all images. Image analysis and preparation of figures were performed in QuPath (25) and FIJI/ImageJ (freely available from NIH). No post-exposure adjustments were made to the images.

### Single-cell transcriptomics analysis

To assess *CALM1-3* gene expression in prenatal brain, we analyzed the single-cell gene expression profile of the entire left hippocampus during gestational weeks 16-27 (26). The data is accessible in the Gene Expression Omnibus (GEO) under accession number GSE131258. Data processing follows the guidelines described (26). Briefly, the gene-cell data matrix was obtained from GEO website. The most recent version of Seurat package (v4.3.0) was used for data analysis. Poor-quality cells with expression of fewer than 800 genes and more than 7000 genes were excluded from the dataset. The genes expressed in at least 30 single cells were entered in the analysis. Hemoglobin-expressing cells and those with more than 15% mitochondrial gene expression were also eliminated. In total, 42954 single cells expressing 17737 genes were included in the downstream analysis. The data underwent normalization for sequencing depth to achieve a total of 10^4^ molecules per cell. Cell types were identified using the top highly variable genes and by the expression of known cell-type markers. Average gene expression levels for the genes of interest were calculated in the Seurat package for all cells in each cluster and for the cells with expression value above zero for the consequent genes for better visualization. The ggplot2 package was used to plot expression patterns across the cell types.

### Calmodulin protein production

Calmodulin variants were expressed in *E. coli* Rosetta 2 (DE3), from a modified pET vector, as fusion proteins carrying an N-terminal maltose-binding protein (MBP) followed by a tobacco etch virus (TEV) cleavage site. Transformed bacteria were grown in LB medium (10 g/l NaCl, 5 g/l yeast extract, 10 g/l tryptone, pH 7.5) at 37°C until reaching the exponential growth phase, then overnight protein expression was induced at 25°C by addition of 1 mM isopropyl β-D-1-thiogalactopyranoside (IPTG).

Bacterial cells were harvested by centrifugation (6000 x g, 10 mins) and lysed by sonication followed by another centrifugation step (40,000 x g, 45 mins) to obtain the lysate. The lysate was filtered (0.2 µm) and applied to a custom packed amylose column (New England Biolabs) attached to an NGC Chromatography System (Bio-Rad). After sample application, the column was washed with three column volumes (CV) of buffer A (20 mM Tris, 100 mM NaCl, 1 mM EDTA, 1 mM DTT pH 7.4) after which the protein was step eluted with 100% buffer B (buffer A added 10 mM maltose).

The MBP-calmodulin fusion protein was cleaved using TEV protease (1:100 w/w) overnight at 4°C. To separate MBP from calmodulin, the sample was applied to a 20 ml Q sepharose FF (Cytiva) column attached to an NGC Chromatography System (Bio-Rad). After sample application, the sample tubing was rinsed with buffer A (20 mM Tris, 100 mM NaCl, pH 7.4), before washing the column with 4 CV 35% buffer B (20 mM Tris, 500 mM NaCl, pH 7.4). Calmodulin was step eluted with 70% buffer B.

The calmodulin-containing fractions were pooled and concentrated to ∼2.5 ml using a 10,000 molecular weight cut-off centrifugal filter in preparation for a final size exclusion chromatography (SEC) step. After concentration, 20 mM EDTA was added to the sample to remove residual calcium ions before applying the sample to a HiLoad 16/600 Superdex 75 pg column (Cytiva) attached to an ÄKTA Purifier chromatography system. The protein was eluted in HK buffer (20 mM HEPES, 100 mM KCl, pH 7.2). The purified calmodulin protein was aliquoted and stored at -80°C until further analysis.

### Calcium/magnesium-buffered solutions

The free calcium concentration was controlled using a dual-chelator buffer system (50 mM HEPES, 100 mM KCl, 2 mM NTA, 0.5 mM EGTA, pH 7.2) (27). Practically, three individual buffers were prepared, all with the previously mentioned composition of chemicals at 1.5x their final concentration. Additionally, one of the three buffers were supplemented with 3 mM calcium (1x), and another was supplemented with 30 mM magnesium (1x). The three buffers (no calcium, high calcium, and high magnesium) were mixed in different ratios to obtain a free magnesium concentration of 1 mM and range of desired free calcium concentrations, calculated using a pCa calculator (27). For calcium titration experiments, a fourth buffer was prepared by spiking the 3 mM calcium buffer to 12.5 mM (1x) with 1M CaCl_2_.

### Calcium binding to calmodulin

Calcium/magnesium buffers (see above) were mixed to obtain a free magnesium concentration of 1 mM and a range of free calcium concentrations, from 40 nM to 8 mM, using an automated Microlab STARlet liquid handling robot (Hamilton). Calmodulin (15 µM) and TCEP (300 µM, 20x [calmodulin]) was added during dilution of the buffers to 1x.

16 discrete calcium/calmodulin solutions were prepared and sequentially transferred to a quartz cuvette. Changes in the intrinsic phenylalanine and tyrosine fluorescence, reflecting calcium binding to the calmodulin N-lobe and C-lobe, respectively, were monitored using a FluoroMax-4 spectrofluorometer (Horiba scientific). For phenylalanine fluorescence, the excitation wavelength was 250 nm (slit 8 nm) and the emission wavelength was 280 nm (slit 8 nm). For tyrosine fluorescence, the excitation wavelength was 277 nm (slit 5 nm) and the emission wavelength was 310 nm (slit 5 nm). Three (tyrosine) or four (phenylalanine) technical recordings were averaged for at least three individual experiments.

The fluorescence intensity (FI) was plotted as a function of the free calcium concentration ([Ca^2+^_free_]) to generate a calcium binding curve. A generic Hill model, taking into account cooperative calcium binding via the Hill coefficient, h, was fitted to determine the apparent calcium binding affinity of calmodulin via the dissociation constant, K_D,app_:

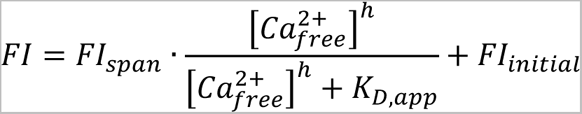

Non-linear curve fitting, according to this model, was done using GraphPad Prism. The dissociation constants were log_10_-transformed before statistical comparison using one-way ANOVA with Dunnett’s *post hoc* test for multiple comparisons.

### Secondary structure determination by circular dichroism

Circular dichroism spectra were recorded at 25°C using a Chirascan Plus spectrometer (Applied Photophysics) for 20 µM calmodulin in a buffer consisting of 4 mM HEPES, 20 mM KCl, 100 µM TCEP, and either 1 mM EDTA or 1 mM CaCl_2_. Spectra were recorded at 190-260 nm with a bandwidth of 1 nm, a step size of 0.5 nm, and a sampling time-per-point of 0.5 seconds. Each spectrum represents the average of two technical recordings for at least four individual measurements. After data collection, each spectrum was buffer subtracted and the signal was converted to mean residue ellipticity, θ_MRE_.

As a spectral descriptor of structural changes caused by calmodulin variants, we calculated the θ signal ratio at 222 and 208 nm, θ_222/208_, reflecting changes in the structure and environment of alpha-helices (highly relevant for calmodulin which consists primarily of alpha-helices both in its apo- and calcium-bound state). Furthermore, as a spectral descriptor of structural changes in the transition from the apo to calcium-bound form, we calculated the change in θ signal at 222 nm, by subtracting the calcium-spectrum signal from the apo-spectrum signal, and dividing it by the apo-spectrum signal, Δθ_222_/θ_222_. Statistical testing for structural differences from the WT was done using one-way ANOVA and Dunnett’s *post hoc* test for multiple comparisons.

### Calcium-dependent calmodulin binding to the Ca_V_1.2 IQ-domain

The interaction between calmodulin and the IQ-domain from Ca_V_1.2 was monitored by changes in the fluorescence anisotropy (FA) of a 5-TAMRA-labeled human Ca_V_1.2-IQ peptide (DEVTVGKFYATFLIQEYFRK-FKKRKEQGLVGKPS) during titration with calmodulin (28). Calcium/magnesium buffers (see above) were mixed to obtain a free magnesium concentration of 1 mM and a range of free calcium concentrations, from 3 nM to 400 µM, using an automated Microlab STARlet liquid handling robot (Hamilton). The TAMRA-labeled peptide (∼20 nM) was added during dilution of the buffers to 1x. Using the liquid handling robot, 3 µl calmodulin was added to the first column of a 384-well plate (Corning) followed by 65 µl calcium buffer/peptide solution. For the four lowest calcium concentrations, a ∼600 µM calmodulin stock was used, whereas for the four highest calcium concentrations, a 22.7 µM calmodulin stock was used. Then, 30 µl calcium buffer/peptide solution was aliquoted into the remaining wells in the plate, resulting in one specific calcium concentration per row. Finally, calmodulin was serial diluted by column-wise transferring 38 µl liquid and discarding the last 38 µl. Half a 384-well plate was used for each calmodulin variant, resulting in eight calmodulin binding curves at eight different calcium concentrations. The FA signal was measured at 25°C in a Spark plate reader (TECAN) using a 535 (25) nm excitation filter and a 590 (20) nm emission filter, a 560 dichroic mirror, 20 flashes, a 40 µs integration time, a 200 ms settling time, a gain of 82, and a z-height of 19740 µm. A G-factor of 0.99 was determined using free 5-TAMRA.

### For each of the eight binding curves, a stoichiometric binding model was fitted

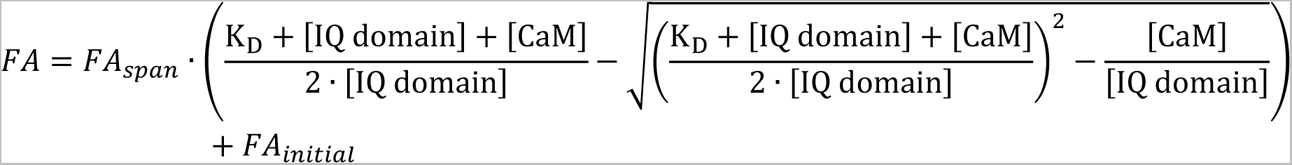

Where K_D_ is the dissociation constant, and [IQ domain] and [CaM] is the total concentration of the 5-TAMRA-labeled IQ peptide and calmodulin, respectively. Non-linear curve fitting, according to this model, was done using GraphPad Prism v6.07. At least three independent experiments were performed for each calmodulin variant. The dissociation constants were log_10_-transformed before statistical comparison using one-way ANOVA with Dunnett’s *post hoc* test for multiple comparisons at each of the eight calcium concentrations.

A plot of the log_10_ transformed K_D_-values as a function of the log_10_ transformed free calcium concentration resulted in a sigmoidal calcium-dependent affinity curve. As a measure of the calcium sensitivity of the calmodulin:Ca_V_1.2-IQ interaction, an variable slope EC_50_ model was fitted to these curves:

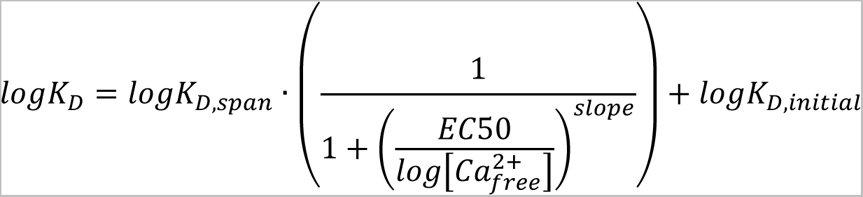

Statistical comparison of EC_50_ values was done using one-way ANOVA with Dunnett’s *post hoc* test for multiple comparisons.

### Preparation of HEK293 Cells for Whole-Cell Patch Clamp

For electrophysiological experiments in HEK293 cells, the α_1C_ plasmid encoding a human channel variant containing exon 8a, within a pcDNA3 expression vector (17) was employed (Acc# Z34810). The WT CaM construct was the human *CALM1* cDNA (10) within the pIRES2-EGFP vector (Clontech Laboratories). From this WT CaM-pIRES construct, pathogenic CaM mutations were generated using QuikChange Lightning™ site-directed mutagenesis (Agilent). HEK293 cells were cultured in 10-cm dishes on glass coverslips, and α_1C_ channels were transfected using an established calcium phosphate method (29). 8 μg each of human α_1C_ cDNA, rat brain β_2a_ (M_80545), rat brain α_2_δ (NM_012919.3) subunits, and WT or mutant CaMs were heterologously co-expressed. Expression was further increased with 2 μg of simian virus 40 T antigen (Tag) cDNA co-transfected. Expression of all constructs was driven by a cytomegalovirus (CMV) promoter.

### Electrophysiological Preparations

Whole-cell voltage-clamp recordings of HEK293 cells were carried out 24-48 hours post-transfection at ambient room temperature. An Axopatch 200B amplifier (Axon Instruments) was used to obtain recordings. These recordings were lowpass filtered at 2 kHz and digitally sampled at 10 kHz to minimize noise. Series resistances of 1.5-4 MΩ were utilized, as well as P/8 leak subtraction. Internal solutions contained (in mM): CsMeSO_3_, 114; CsCl, 5; MgCl_2_, 1; MgATP, 4; HEPES (pH 7.4), 10; and BAPTA, 10; at 295 mOsm adjusted with CsMeSO_3_. External solutions contained (in mM): TEA-MeSO_3_, 140; HEPES (pH 7.4), 10; and CaCl_2_ or BaCl_2_, 40; at 300 mOsm, adjusted with TEA-MeSO_3_. These solutions produced the following uncorrected junction potentials: 10 BAPTA/40 Ca^2+^: 10.5 mV; 10 BAPTA/40 Ba^2+^: 10.2 mV (30). The fraction of peak current after a 300-ms step depolarization (*r_300_*) at test voltages was measured. Ca^2+^/CaM-dependent inactivation (CDI) was evaluated as *f_300_* = (Ba *r_300_* – Ca *r_300_*).

### Statistical analyses

Statistical analysis was performed with GraphPad Prism or Mathworks MATLAB using indicated methods. The significance levels are given as: *p<0.05, **p<0.01, ***p<0.001, ****p<0.0001. Logistic regression was used to test for association between calmodulin missense carrier status and schizophrenia disease status. The association test was based on the asymptotic assumption that the likelihood ratio test (*LR* = 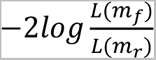), where 𝐿(𝑚_*f*_) is the likelihood of the full model including the variant status, while 𝐿(𝑚_*r*_) is the likelihood of the reduced model neglecting the variant status) follows a 𝜒^2^ distribution with one degree of freedom (31).

### Preparation of plots and figures

Data handling was performed using Microsoft Excel, GraphPad Prism, R, and Mathworks MATLAB. Microscope images were prepared in QuPath and FIJI ImageJ. Figures were prepared in GraphPad Prism, Mathworks MATLAB, and Inkscape.

## Results

### Calmodulin C-lobe variants associate with increased SCZ risk

The Schizophrenia Exome Sequencing Meta-Analysis (SCHEMA) contains exomes from 24,248 patients with schizophrenia or schizoaffective disorder and from 97,322 control individuals with no reported psychiatric disorder (23). In this collection, we found in total 27 carriers of missense variants in *CALM1-3* (Fig 1, Table S1). Seven of these carriers were patients with schizophrenia (SCZ) (carrier frequency ∼1:3,500), while the remaining 20 variants were observed in the control population (carrier frequency ∼1:4,900).

**Figure 1:**
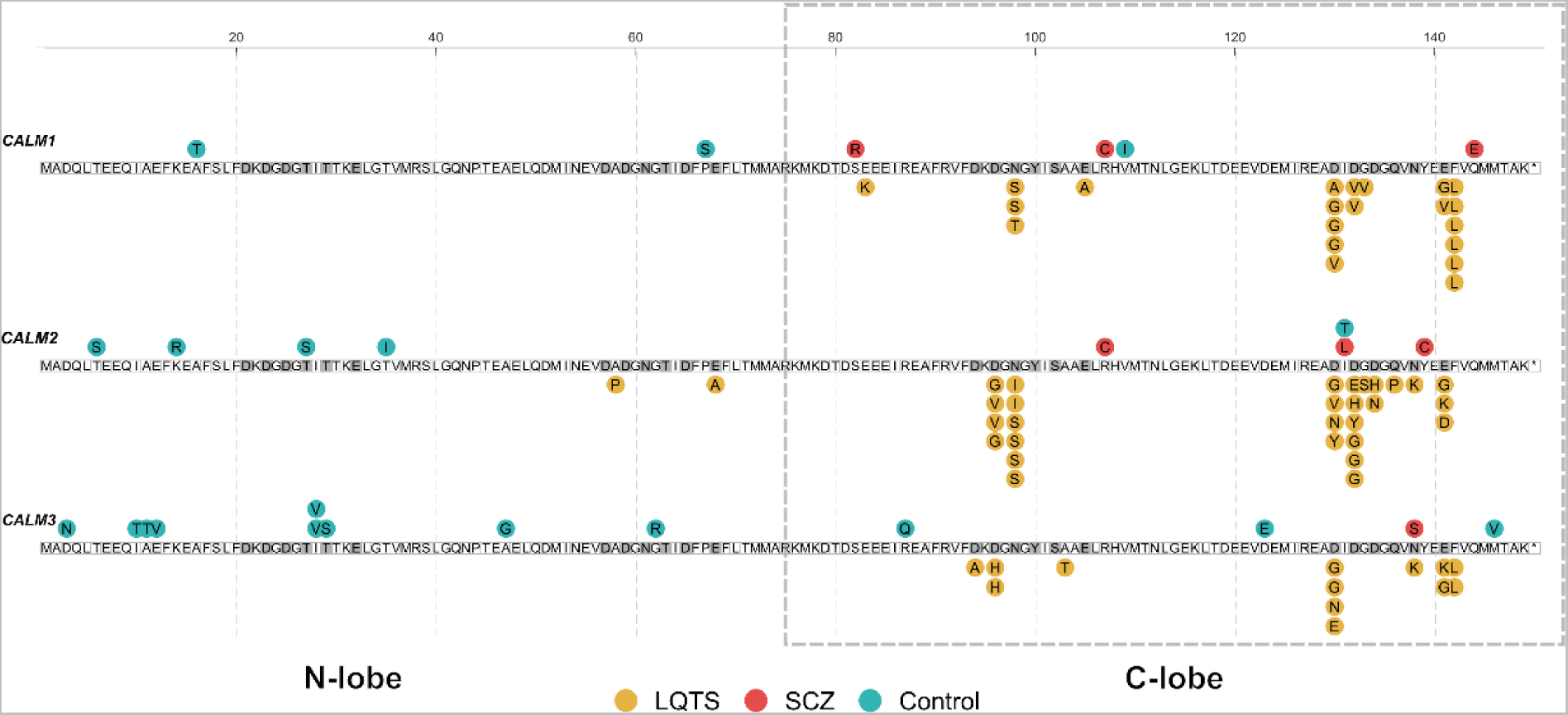
*CALM1-3* variants in LQTS and SCZ patients occur in the calmodulin C-lobe. Three genes, *CALM1-3*, produce identical calmodulin proteins, represented by the three rows. Numbering follows the immature 149 amino acid sequence, although the initial methionine is cleaved off during translation. Each circle represents an independent case of a calmodulin variant carrier with LQTS (yellow), SCZ (red), or control carriers (teal), with the associated amino acid position and substitution indicated. The C-terminal lobe of calmodulin, harboring the majority of LQTS and all SCZ variants, is framed with a dashed square. Amino acids that coordinate calcium are highlighted in dark gray. *LQTS: Long QT syndrome. SCZ: schizophrenia*.

We first calculated the odds ratio (OR) for SCZ, being carrier of a missense variant in any of the calmodulin genes. Here, we found no statistically significant effect (OR 1.4±0.62, *P_adj_*-value 0.57) (Table 1). Because the two domains in calmodulin are functionally different, we tested if the variants were equally distributed between the two lobes in SCZ cases and controls. We noted that all seven variants from SCZ patients were exclusively observed in the calmodulin C-lobe, while there were five C-lobe variants and 15 N-lobe variants in the control population (Fig 1). This remarkable separation (0 SCZ and 15 control variants in N-lobe; 7 SCZ and 5 control variants in C-lobe) was statistically significant (*P* = 0.0009, Fisher’s Exact test). Thus, we determined the OR for SCZ, carrying a variant in the C-lobe. Here, we found an increased risk (OR 5.6±3.3, *P_adj_*-value 0.043).

**Table 1:** Odds ratio calculations for *CALM1-3* variants in SCZ cases and controls. * False discovery rate (FDR)-adjusted *P*-values. *OR, odds ratio, SE, standard error*.

|  | SCZ<br>*CALM* carriers<br>(n = 24248) | Control<br>*CALM* carriers<br>(n = 97322) | OR | SE | P-value | *P*<sub>adj</sub>-value* |
| --- | --- | --- | --- | --- | --- | --- |
| ***CALM1+2+3*** | 7 | 20 | 1.40 | 0.62 | 0.52 | 0.57 |
| N-lobe | 0 | 15 | 3.8×10<sup>-5</sup> | 1.9×10<sup>-3</sup> | 0.0098 | 0.054 |
| C-lobe | 7 | 5 | 5.62 | 3.29 | 0.0036 | 0.043 |
| ***CALM1*** | 3 | 3 | 4.01 | 3.28 | 0.10 | 0.20 |
| N-lobe | 0 | 2 | 2.8×10<sup>-4</sup> | 0.014 | 0.35 | 0.42 |
| C-lobe | 3 | 1 | 12.0 | 13.9 | 0.018 | 0.054 |
| ***CALM2*** | 3 | 5 | 2.4 | 1.8 | 0.25 | 0.33 |
| N-lobe | 0 | 4 | 2.8×10<sup>-4</sup> | 0.010 | 0.18 | 0.31 |
| C-lobe | 3 | 1 | 12.0 | 13.9 | 0.018 | 0.054 |
| ***CALM3*** | 1 | 12 | 0.33 | 0.35 | 0.22 | 0.33 |
| N-lobe | 0 | 9 | 1.0×10<sup>-4</sup> | 0.0041 | 0.045 | 0.11 |
| C-lobe | 1 | 3 | 1.3 | 1.5 | 0.81 | 0.81 |

The pathogenicity of genetic variants can be predicted using different computational tools. The SCHEMA study uses the MPC pathogenicity score (32), where MPC≥2 is considered pathogenic. Of the 27 calmodulin variants, eight have an MPC score ≥2. Of these eight variants, four are in the C-lobe in SCZ patients, and four are in the N-lobe in control individuals (Table S1). When restricting to variants with MPC score ≥2, the association of C-lobe variants with SCZ has the same effect size but the *P*-value becomes lower despite fewer variants (OR 5.7×10^4^±2.1×10^6^, *P_adj_*-value 0.0036), suggesting that the association signal comes from variants with MPC≥2 (Table S2).

We also noted a different distribution of SCZ and control variants between the three genes (Fig 1). Of the seven variants in SCZ patients, three are in *CALM1*, three in *CALM2*, and one in *CALM3*. On the contrary, of the five C-lobe variants in the control group, one is in *CALM1*, one in *CALM2*, and three in *CALM3*. Therefore, we evaluated the contribution of each *CALM* gene to the increased SCZ risk associated with calmodulin C-lobe variants (Table 1). Although not significantly different, C-lobe variants result in a different pattern of risk association for *CALM1* and *CALM2* (both OR 12.0±13.9, *P_adj_*-value 0.054) compared to *CALM3* (OR 1.3±1.5, *P_adj_*-value 0.81).

Together, these observations demonstrate an increased SCZ risk from calmodulin C-lobe variants and indicate that most of this risk comes from *CALM1* and *CALM2*. Of note, calmodulin variants in LQTS patients mainly affect the C-lobe (7), emphasizing the importance of considering the two lobes separately when assessing effects of missense variants. It is also worth noting that we find no identical variants between LQTS patients, SCZ patients, and control individuals.

### *CALM1*, *CALM2*, and *CALM3* are highly expressed in the brain

Calmodulin protein levels are high in the brain (∼0.5mg/g) (3), but since the protein product of the *CALM1*, *CALM2*, and *CALM3* genes is identical, it cannot be easily determined if the protein is produced from all three genes. Because the variant pattern in *CALM3* stood out from that in *CALM1* and *CALM2*, we investigated if all three genes are expressed in the brain. To study the *CALM* expression level in different brain regions, we used the Genotype-Tissue Expression (GTEx) database (24). For comparison, we included heart left ventricle, skeletal muscle, and lung tissues. As a low expression control, we included *CLDN4*, which encodes claudin 4 and is primarily expressed in epithelia, including the lung (33) (Fig 2A, first row). As a high expression control, we included *UBC*, which encodes polyubiquitin C and is highly expressed in all tissues (34, 35) (Fig 2A, second row). For reference, we also included *CACNA1C* and ten other genes (*CACNA1G, CUL1, GRIA3, GRIN2A, HERC1, RB1CC1, SETD1A, SP4, TRIO, XPO7*), for which rare genetic variants have been associated with increased SCZ risk in the SCHEMA study (23). Strikingly, all three *CALM* genes had the highest expression level across all brain tissues and genes investigated, even higher than that of the *UBC* positive control (Fig 2A).

**Figure 2:**
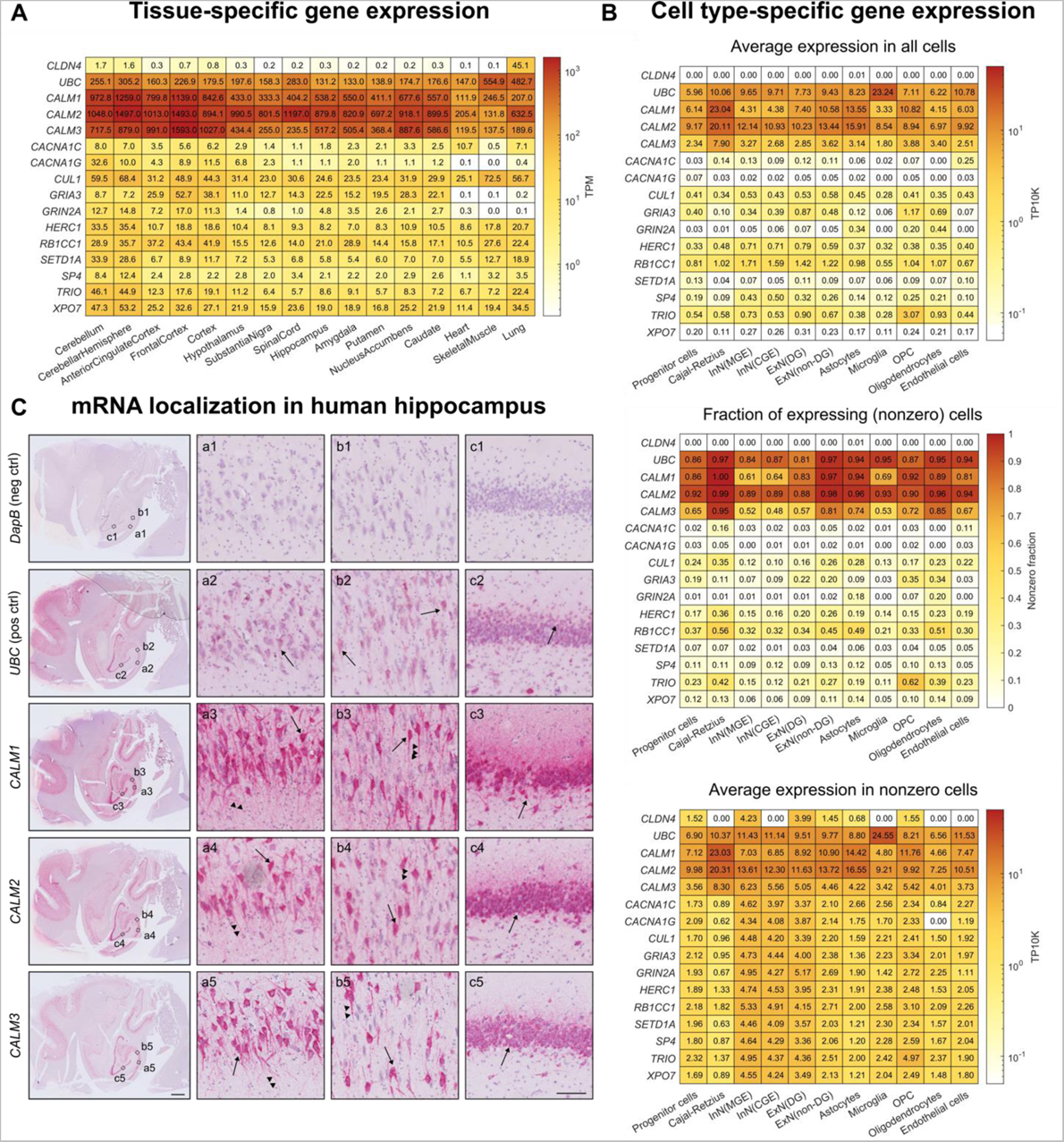
High levels of *CALM1-3* mRNA in the brain. **A-B**. mRNA levels of *CALM1-3*, *CLDN4* (negative control), *UBC* (positive control), *CACNA1C* (encodes Ca_V_1.2), and ten genes for which rare genetic variants are associated with increased SCZ risk (23). **A.** Transcript levels extracted from the genotype-tissue expression (GTEx) project. **B.** Single-cell spatial transcriptomics data from human hippocampal fetuses at gestational weeks 16-27 (26) (see also Fig S1-3). **C.** *In situ* hybridization with RNAscope probes (red signal) targeting the indicated genes of a healthy human donor. Arrows indicate examples of positively stained neuronal cell bodies, and arrowheads indicate examples of stained neuronal processes. The images are representative of three individual stains of two donors (see also Fig S4). Scale bars are 2 mm and 100 µm for insets. *ExN(DG): Dentate gyrus excitatory neurons. ExN(non-DG): Non-dentate gyrus excitatory neurons. InN(CGE): Caudal ganglion eminence-derived inhibitory neurons. InN(MGE): Medial ganglion eminence-derived inhibitory neurons. OPC: oligodendrocyte progenitor cells. TP10K: Transcripts per 10,000. TPM: Transcripts per million*.

We next zoomed in to further explore the expression patterns of these genes in a single-cell spatial transcriptomics sequencing dataset from human fetal hippocampus (26). Supporting our findings on the tissue-level, *CALM1*, *CALM2*, and *CALM3* expression levels were high and similar to the positive control across all hippocampal cell types (Fig 2B, upper panel) and across all gestational stages (Fig S1-3). During analysis, we noticed that this dataset – like other spatial transcriptomics datasets – included a subset of cells with zero *CALM* gene expression (36) (Fig S1-3). It is unclear whether the presence of *CALM*-zero cells is a biological finding or a technical error (36). Therefore, we determined the expression levels in the non-zero cells population only (Fig 2B, middle and bottom panels). First, we calculated the fraction of non-zero-to-total number of cells and found that *CALM1* and *CALM2* expression was measured in 61-100% of the cells, whereas *CALM3* expression was measured in 48-95% (Fig 2B, middle panel). When considering expression levels only in non-zero cells, we consistently found that all three *CALM* genes were expressed across the brain at levels similar to or higher than the included controls and references (Fig 2C, lower panel).

These results were corroborated by *in situ* hybridization of *CALM1*, *CALM2*, and *CALM3* in adult human hippocampus. We stained hippocampal tissue from two healthy donors using specific RNAscope probes directed at *CALM1*, *CALM2*, or *CALM3* mRNAs (Fig 2C, Fig S4). Consistent with the expression results presented above, we observed that all three *CALM* genes were highly expressed across the tissue, including in pyramidal cells in the CA regions (Fig 2C, a1-b5) and in granule cells in the dentate gyrus (Fig 2C, c1-5).

Together, all three *CALM* genes were thus consistently highly expressed in the brain, across all examined regions, particularly in neurons. Therefore, although *CALM3* stands out in the statistical risk estimations, we included variants from all three *CALM* genes in subsequent *in vitro* studies.

### SCZ calmodulin variants affect calcium binding affinity

The primary function of calmodulin is to bind calcium ions (Fig 3A). Therefore, we next asked if variants in SCZ patients and control individuals affect the calcium binding affinity of calmodulin. We included all SCZ variants (six unique variants at the protein level from seven carriers). From the control group, we included all five variants in the C-lobe and nine unique protein variants from ten carriers in the N-lobe. We asked if the variants affected the calcium binding affinity of the lobe they are located in.

**Figure 3:**
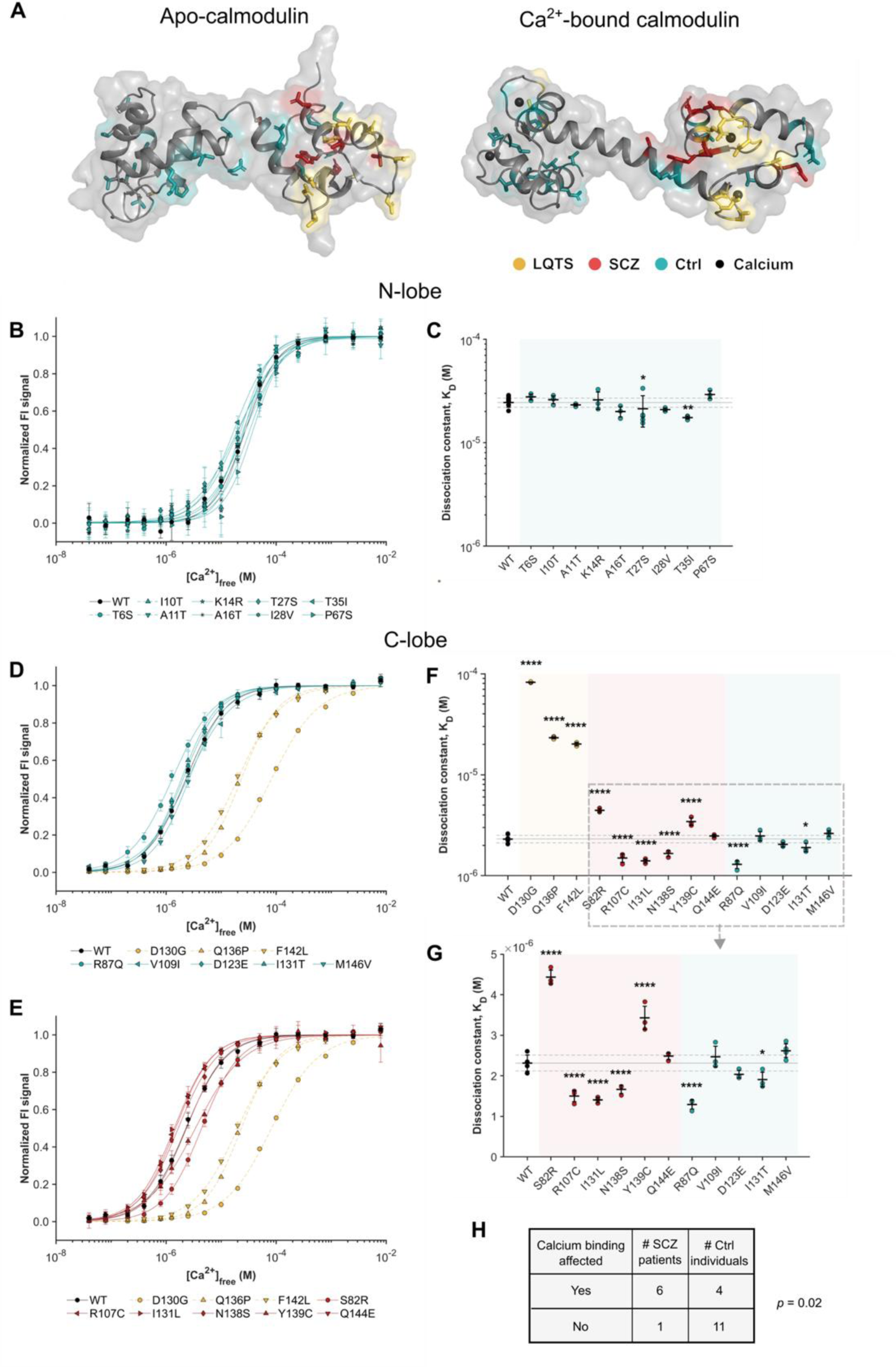
Calmodulin SCZ variants significantly affect calcium binding. **A.** Visualization of calmodulin amino acid positions affected in LQTS patients (yellow), SCZ patients (red), and in control individuals (teal) on the crystal structure of calmodulin (gray) in apo-state (PDB: 1DMO) or with calcium ions (black) bound (PDB: 1CLL). **B.** Calcium binding curves of the indicated N-lobe calmodulin variants. **C.** Calcium affinities of the indicated N-lobe calmodulin variants. **D-E.** Calcium binding curves of the indicated calmodulin variants in C-lobe. Data normalization in B, D, and E was only done for plotting purposes. All fitting was done on raw data. **F-G.** Calcium affinities of the indicated C-lobe calmodulin variants. **H.** Summarized effect on calcium binding in SCHEMA carriers of calmodulin variants. Statistics: Fisher’s exact test. *K_D_, dissociation constant*.

To determine the calcium affinity of each variant, we used a well-established assay in which calcium binding is monitored by the intrinsic phenylalanine (N-lobe) or tyrosine (C-lobe) fluorescence of the protein (Fig 3B-G) (37–39). From a generic Hill fit, the calcium binding affinity can be determined, represented by the dissociation constant, K_D_ (Fig 3B,D,E, Table S3,S4). An increase in the K_D_ value reflects decreased binding affinity. For comparison, we included three LQTS-causing variants known to impair calmodulin’s calcium affinity and thereby cause a loss-of-function effect (11, 12): D130G, Q136P, and F142L (Fig3D-F). These arrhythmogenic variants reduced the C-lobe calcium affinity by 8-35-fold, compared to the WT. In comparison, all SCHEMA variants changed the calcium affinity by less than 2-fold, regardless of lobe position (Fig3B-G, Table S3,S4).

Interestingly, we found significant differences of calmodulin variants in both the SCZ and control group (Fig 3B-G). Five of the six variants from SCZ patients significantly affected the calcium affinity: S82R and Y139C reduced the calcium binding affinity by 1.92- and 1.48-fold, respectively, compared to the WT – a pattern similar to that of arrhythmogenic variants, despite the lower magnitude. In contrast, and surprisingly, R107C, I131L, and N138S increased the calcium affinity by 1.39-1.54-fold (Fig 3D, Table S4). Similarly, four variants from control group, T27S, T35I, R87Q and I131T, increased the affinity by 1.15-1.79-fold, compared to the WT (Fig 3D, Table S3-4). Notably, an increased calcium affinity, a gain-of-function effect, has never before been reported for any human calmodulin variant.

To understand if variants with calcium binding effects were differently distributed between SCZ patients and control individuals, we quantified the number of carriers where calmodulin variants displayed significant calcium binding effects (Fig3H). We found that this simple measure – if a variant significantly affects calmodulin calcium sensing – was enough to statistically separate the two groups. Thus, calmodulin variants in SCZ patients are more likely to affect calmodulin calcium sensing, suggesting a close link between the effect of the variant on protein function and SCZ.

### SCHEMA calmodulin variants perturb the apo-structure of calmodulin

Since we found a significant enrichment of calcium binding effects in SCZ patients compared to control individuals, we asked whether the changes in calcium affinity reflected a variant-induced structural change in calmodulin. To this end, we recorded circular dichroism spectra both in the apo-state and the calcium-bound state (Fig S5-6). We included all SCZ variants, all C-lobe control variants, and the three LQTS variants D130G, Q136P, and F142L. Since calmodulin primarily consists of α-helices in both its forms, we extracted the signal intensity at 222 and 208 nm and calculated the intensity ratio 222/208; a spectral descriptor that reflects the α-helical conformation and environment (40, 41) (Fig 4A-B, Table S5). To describe the change in α-helical conformation between the apo and calcium-bound state, we determined the change in signal intensity at 222 nm between the two conditions (Fig 4C, Table S5).

**Figure 4.**
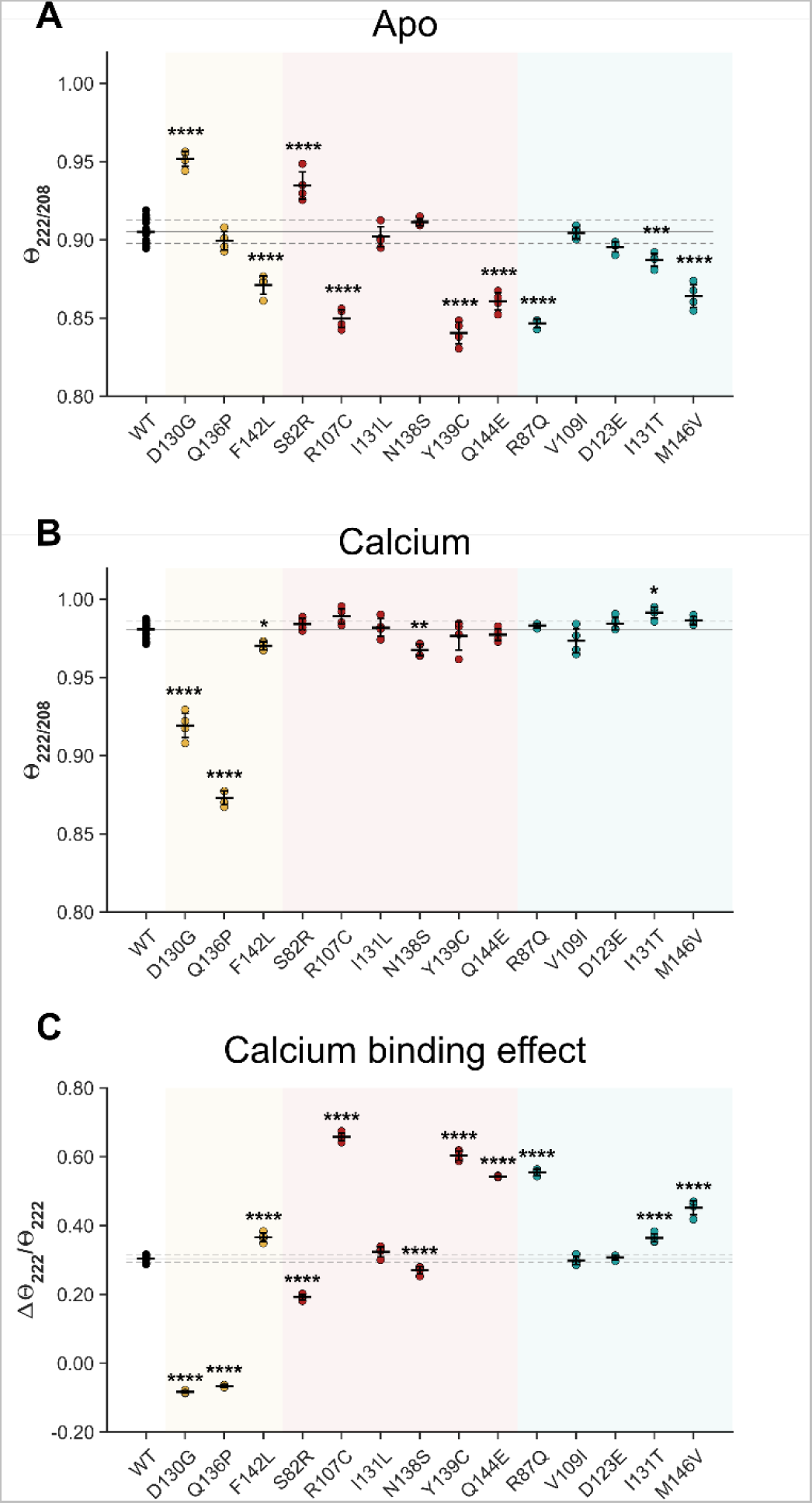
Calmodulin variants in SCHEMA primarily affect the apo structure. Circular dichroism was used to determine structural effects of the indicated variants on the α-helixes of apo-calmodulin (A) or calcium-bound (B) as well as the calcium-induced difference between the two states (C). Data for D130G and Q136P is reproduced from a previous study (42). Raw spectra are shown in Fig S5-6, and summarizing statistics are in Table S5.

Among the three LQTS-variants, we found that D130G and F142L, but not Q136P, significantly affected the structure of apo-calmodulin (Fig 4A). On the other hand, D130G and Q136P, but not F142L, affected the structure of calcium-calmodulin (Fig 4B). Thus, although the calcium affinity is impaired in all three cases, stand-alone structural changes do not necessarily reflect this change and suggest different underlying biophysical mechanisms, likely reflecting the different roles for the affected residues, e.g. if they directly coordinate calcium or not.

For the SCHEMA variants, we found that SCZ variants S82R, R107C, Y139C, and Q144E, and control variants R87Q, I131T, and M146V, had significant structural effects on the apo-form (Fig 4A). In the calcium-bound form, only SCZ variant N138S and control variant I131T showed significant effects (Fig 4B). Noteworthy, while the structural effects of the SCHEMA variants were much smaller than the effects of the LQTS variants on the calcium-bound form, they were similar in magnitude to those of the LQTS variants in the apo-form. Considering the change in α-helical content between apo and calcium-bound form, we found that the SCZ variants S82R, R107C, N138S, Y139C, and Q144E and the control variants R87Q, I131T, and M146V had significant effects (Fig 4C).

Overall, these structural data demonstrate that while LQTS-variants affect both apo- and calcium-bound calmodulin, the variants found in the SCHEMA cohort primarily affect apo-calmodulin.

### SCZ calmodulin variants perturb calcium sensitivity during Ca_V_1.2-IQ binding

The pathology of calmodulin variants in LQTS is largely linked to dysregulation of Ca_V_1.2 (10). Since Ca_V_1.2 variants are also implicated in SCZ, we asked if calmodulin variants from SCZ patients and control individuals affect Ca_V_1.2 binding and regulation. To answer this question, we monitored the interaction between calmodulin and the IQ-domain of Ca_V_1.2 at calcium concentrations spanning resting and activating cellular conditions (28) (Fig S7-9). Across this range of calcium concentrations, the binding affinity of calmodulin towards Ca_V_1.2-IQ increases by more than 10,000 fold, emphasizing the sophisticated sensor function of calmodulin. To measure this calcium-dependent change in affinity, we determined the dissociation constant, K_D,_ of the calmodulin:Ca_V_1.2-IQ complex at each calcium concentration (Fig 5A-B).

**Figure 5:**
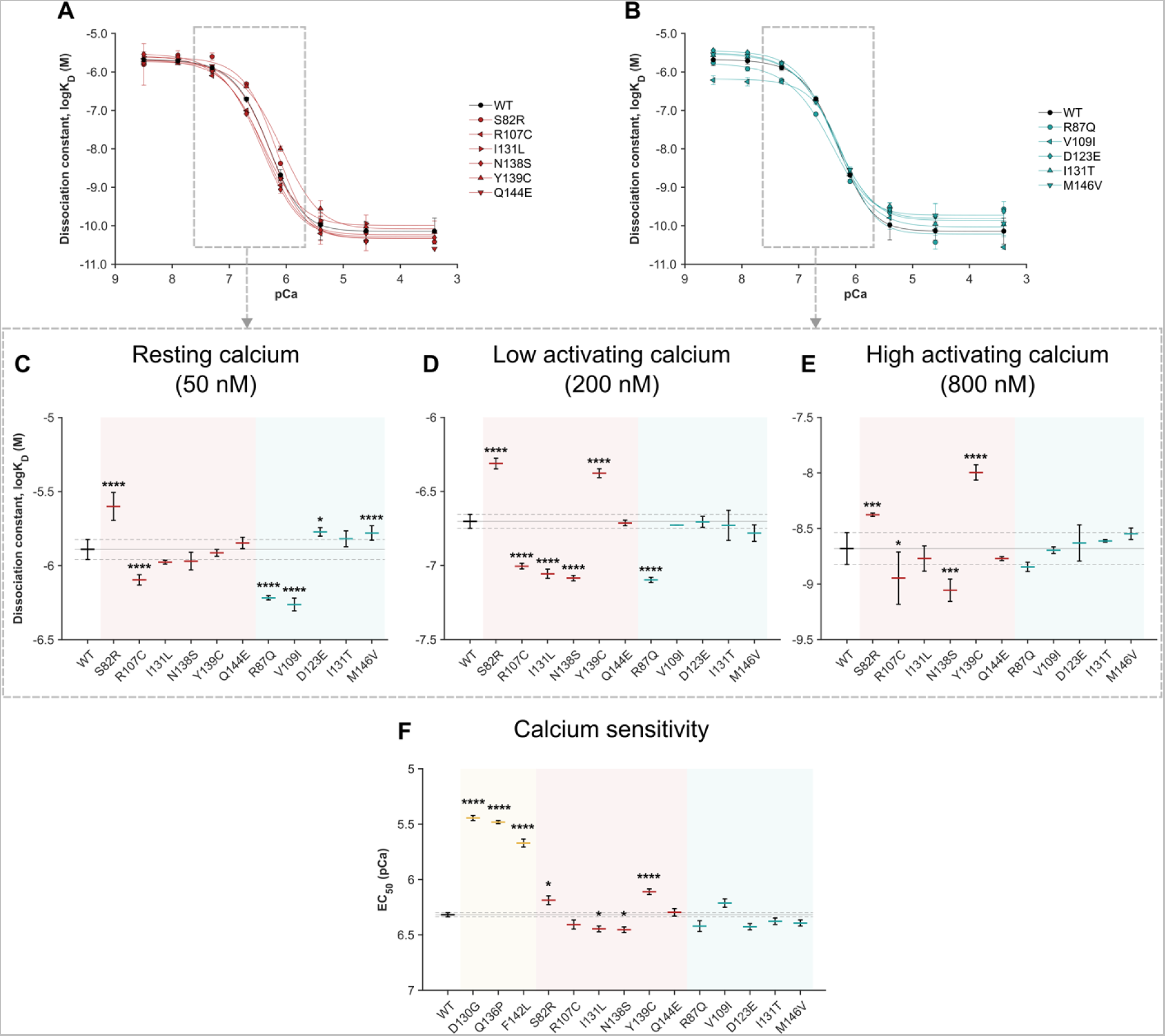
Calmodulin SCHEMA variants have differential effects on calcium-dependent Ca_V_1.2-IQ binding. **A-B.** Binding affinity, represented by the dissociation constant, K_D_, of calmodulin binding to the IQ peptide of Ca_V_1.2 as a function of free calcium concentration. Dashed squares indicate data presented in detail in C-E. **C-E**. Calmodulin:Ca_V_1.2-IQ affinity at 50 nM, 200 nM, and 800 nM calcium, representing three different physiological conditions. **F.** EC_50_ values reflecting the calcium sensitivity of calmodulin when bound to the Ca_V_1.2-IQ domain. Data for D130G, Q136P, and F142L is reproduced from previous studies (42, 44). *EC_50_, half maximal effective concentration, Ca_V_1.2, voltage-gated calcium channel 1.2*, *pCa, -log([Ca^2+^])*.

At low resting calcium concentrations (50 nM, Fig 4C) (43), we found that one SCZ variant (S82R) and two control variants (D123E and M146V) caused a significant decrease in the Ca_V_1.2-IQ affinity of calmodulin (loss-of-function), whereas another SCZ variant (R107C) and two other control variants (R87Q and V109I) caused a significant increase in affinity (gain-of-function). Initially in the calcium-induced transition from low to high Ca_V_1.2-IQ affinity (200 nM free calcium, Fig 4D), we found that all SCZ variants, except Q144E, significantly changed the Ca_V_1.2-IQ affinity, albeit in different directions: S82R and Y139C decreased the affinity, whereas R107C, I131L, and N138S increased it. In the control group, one variant (R87Q) significantly increased the affinity. Later in the calcium-induced transition (800 nM free calcium, Fig 4E), four SCZ variants retained their significant effects on the Ca_V_1.2-IQ interaction (S82R, R107C, N138S, and Y139C), whereas the control group variants no longer had any significant effect.

Finally, at high calcium concentrations (> 800 nM) reflecting a Ca^2+^-saturated calmodulin:Ca_V_1.2-IQ complex, the affinity was too high to determine accurately, and statistical testing could thus only be done for a subset of the calmodulin variants. At these calcium concentrations, there were no significant effects of any of the measurable calmodulin variants (Table S6).

As an overall measure of the variant effect on the calcium-dependency of calmodulin binding to Ca_V_1.2-IQ, we determined the EC_50_ from all curves (Fig 5F, Table S7). For comparison, we included already published data for the three LQTS variants D130G, F142L, and Q136P (42, 44). Among the calmodulin variants from SCZ patients, we found that S82R and Y139C significantly increased the EC_50_, suggesting a reduction in the calcium sensitivity of calmodulin in the calmodulin:Ca_V_1.2-IQ complex. On the contrary, SCZ variants I131L and N138S significantly decreased the EC_50_, suggesting an increased calcium sensitivity of calmodulin in the complex. Remarkably, among the calmodulin variants from control individuals, we found no statistical differences from the WT. Of note, compared to the EC_50_ effect of the three calmodulin variants in LQTS patients, the effects in SCZ patients were substantially smaller in magnitude.

The effects of the SCZ variants on Ca_V_1.2-IQ binding affinity and calcium sensitivity correlate excellently with the effects on calcium binding. S82R and Y139C, found to reduce the calcium affinity of calmodulin, impair Ca_V_1.2-IQ binding and calcium sensitivity. In contrast, R107C, I131L, and N138S, found to increase the calcium affinity of calmodulin, enhance Ca_V_1.2-IQ binding and calcium sensitivity.

### The calmodulin variant Y139C impairs Ca_V_1.2 closure

Calmodulin binding to Ca_V_1.2 binding, and subsequent calcium sensing, is translated into calcium dependent inactivation (CDI) of the channel. Since calmodulin variants from SCZ patients, but not control individuals, affected the calcium-sensitivity of calmodulin when bound to Ca_V_1.2-IQ, we asked if these differences translate to an effect on CDI, using whole-cell voltage-clamp recordings of transfected HEK293 cells.

Cells were clamped at defined voltages (Fig 6). Upon clamping, the channels open, and the inward ion current is visualized as a downward deflection of the exemplar current traces (Fig 6A-B, left panels). Shortly after, a rapid decay in calcium current (red or yellow trace) is inflicted by calmodulin in response to sensing calcium, thereby resulting in CDI. For reference, the same experiment is conducted with barium instead of calcium. Calmodulin cannot bind barium and thus does not induce CDI, and the barium current remains at almost the same level during the experiment (black trace). The calcium and barium currents remaining after 300 ms are quantified as *r_300_*, and the difference between the two is calculated as *f_300_* (Fig 6C, Table S8, Fig S10).

**Figure 6:**
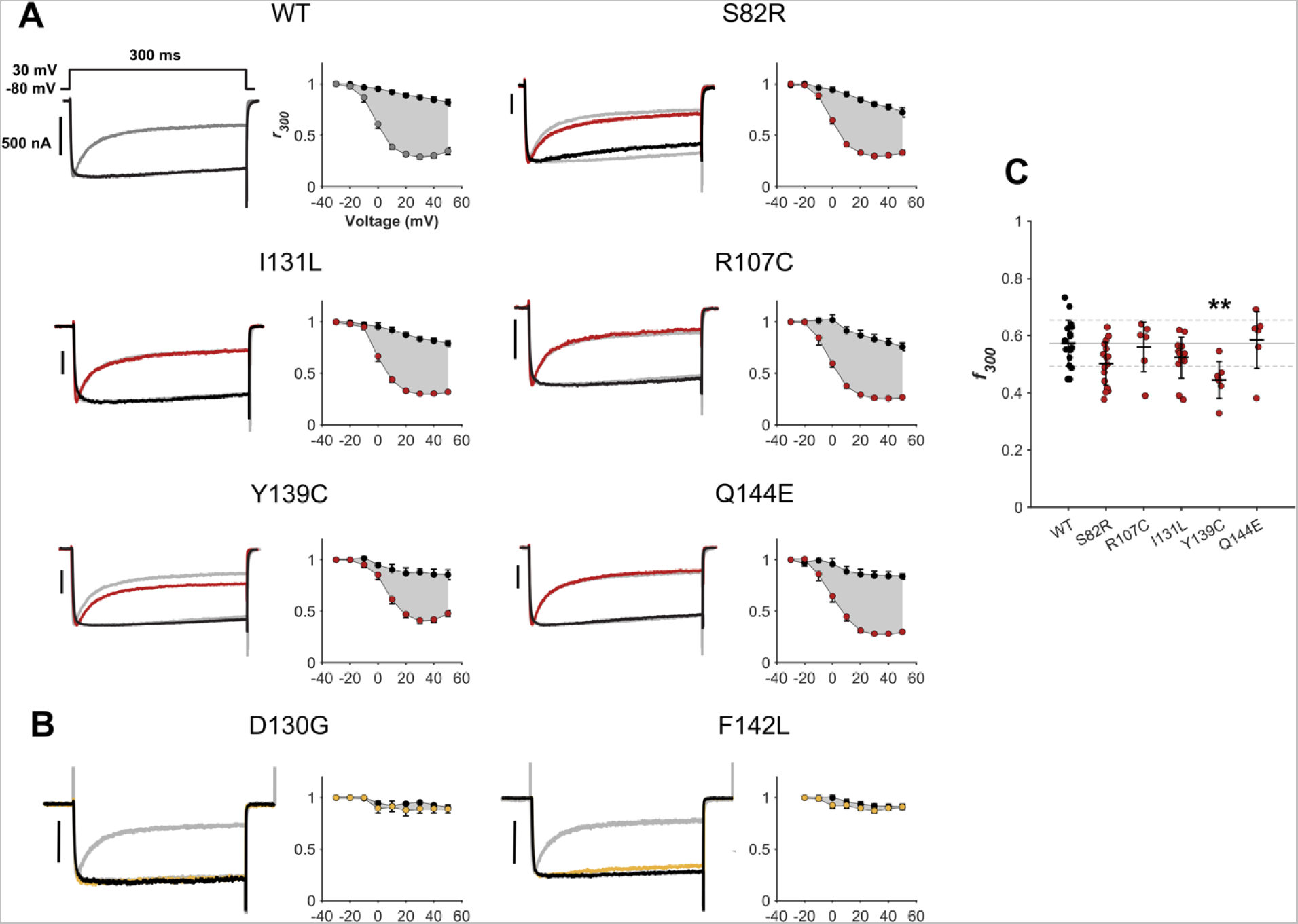
Small effects of calmodulin variants from SCZ patients on CDI of the Ca_V_1.2 channel. **A-B.** Left panels show exemplar currents evoked by 30-mV voltage step in cells co-transfected with Ca_V_1.2 and the indicated calmodulin variants. For comparison, the trace for WT calmodulin is reproduced in gray behind the exemplar traces for the indicated variants. **A.** Calmodulin variants from SCZ patients. **B.** Calmodulin variants from LQTS patients, reproduced from a previous study along with the corresponding WT in gray (10). CDI manifests as the rapid decay in calcium current (colored trace) as compared to barium (black trace). Right panels show population data for CDI across voltages. *R*_300_ measures the current remaining after 300 ms, after normalization to peak current. *F*_300_ is the difference between calcium and barium at 30 mV, after normalization by the barium *r*_300_ value. **C.** Quantification of *f_300_*. Statistical calculations are shown in Table S6.

None of the SCZ variants affected CDI to similar levels as the LQTS variants D130G and F142L (Fig 6A-B). Among the SCZ variants, a significant reduction of 24.8% in *f_300_* was observed for Y139C (0.57±0.08 to 0.445±0.06, *P*=0.007), consistent with a gain-of-function effect on the channel. Although not reaching statistical significance, a small reduction was also observed for S82R (12.5% reduction to 0.50±0.08, *P*=0.074). Interestingly, S82R and Y139C are the only SCZ variants that reduced the calcium affinity of calmodulin and the calcium sensing of calmodulin in the Ca_V_1.2-IQ:calmodulin complex, suggesting that they act through a similar mechanism to that of LQTS patients, but with a smaller impact. Together, these data demonstrate that calmodulin variants from SCZ patients have small or no effects on Ca_V_1.2 CDI.

## Discussion

This is the first study of human calmodulin variants from individuals with no reported cardiac disease. We identified and characterized calmodulin variants from SCZ patients and control individuals in the SCHEMA study. The SCZ calmodulin variants associated with a higher risk of SCZ, and they had a stronger impact on protein function compared to the variants in the control group. These results imply that calmodulin variants can contribute to the development of SCZ.

Our framework for understanding the effects of human calmodulin variants comes from cardiac arrhythmia, primarily LQTS, where variants generally have strong loss-of-function effects on C-lobe calcium affinity and strongly impair Ca_V_1.2 regulation. Reflecting this pattern, all SCZ variants identified in this study occur in the calmodulin C-lobe. However, despite the shared location to the C-lobe, there is no overlap in amino acid substitution between SCZ variants and LQTS variants, and only a single positional overlap on N138 (7). This difference translates into different molecular effects of the two groups of variants: LQTS variants consistently have stronger effects on calcium affinity, protein structure, and Ca_V_1.2 binding and regulation compared to SCZ variants. Moreover, the direction of effect (decreasing or increasing) is the same for LQTS variants in terms of calcium affinity and Ca_V_1.2 binding and regulation, whereas the SCZ variants have differential effects on both protein characteristics. We propose that these differences in effect size determine whether a carrier suffers from LQTS or not.

Although the effects of calmodulin SCZ variants are smaller than LQTS variant effects, they are consistently larger and/or more frequent than those of the control variants, suggesting different molecular effects of the SCZ and control group. Despite the designation ‘control group’, the observation of significant effects among this group of calmodulin variants was not unexpected. First, the calmodulin protein sequence is highly conserved, suggesting a loss-of-fitness from any kind of variation (2). Second, the control population enrolled in the SCHEMA study is a combination of individuals with no known psychiatric disease and individuals from other non-neurological populations (23). Thus, not all controls are directly screened for SCZ, and none are systematically screened for heart disease or other conditions. In principle, these individuals could therefore be at risk of having or developing a psychiatric disorder or other diseases. We do, however, not expect such other phenotypes to be highly detrimental, as that would likely have prevented participation in a genetic screening study as adults. We included calmodulin variants from both SCZ cases and control individuals in the SCHEMA cohort to capture the full allelic spectrum of variants and their functional consequences. Moreover, having such unscreened controls is custom in most genetic association studies.

Along the same lines, the SCHEMA SCZ variants are based on carriers with a known SCZ phenotype, but additional phenotypes could be present and linked to the differential effects observed in this study. Among the SCZ variants, S82R and Y139C have a loss-of-function effect on calcium binding and Ca_V_1.2-IQ binding. Y139C also impacts Ca_V_1.2 closure, whereas S82R has a small, but insignificant effect. Of note, S82R binding to Ca_V_1.2-IQ is significantly reduced under resting calcium conditions. Thus, although overexpressing the protein in the gating experiments, a higher number of channels may be populated by endogenous wildtype protein, masking potential effects on channel regulation. Oppositely, R107C, I131L, and N138S have gain-of-function effects on the calcium and Ca_V_1.2-IQ binding affinity, but have no effects on Ca_V_1.2 regulation in our assay. This is the first time such gain-of-function in binding affinity has been reported for a human calmodulin variant. As these effects do not translate to an overt effect on Ca_V_1.2 regulation, we speculate if the variants could affect other brain targets through an alternative mechanistic paradigm.

Since we identified the first human *CALM* variants, and linked them to cardiac arrhythmia, many such variants have been identified and characterized (1, 7, 15). Because of the link to life-threatening cardiac arrhythmia, mainly LQTS, the search for novel variants, and underlying molecular mechanisms, has primarily involved cardiac patients and phenotypes. Yet, in the recently updated ICalmR (7), 20 of 111 cardiac calmodulinopathy patients also suffer from neurological features such as intellectual disability, seizures, attention-deficit/hyperactivity disorder (ADHD), and autism spectrum disorder (ASD). Moreover, a single patient carrying a I64M variant is reported to have gross neurological and developmental defects, without any cardiac consequences (7). Thus, accumulating evidence suggests that human calmodulin variants can cause or contribute to neurological disease.

This notion is corroborated by genetic studies that suggest a large overlap in the genetic factors underlying neurodevelopmental disorders, including ADHD, ASD, intellectual disability, and SCZ (45). SCZ as well as other neurodevelopmental disorders are highly heritable and large sequencing studies – including SCHEMA – have identified long lists of genetic variants contributing to these diseases. Together, they paint a complex picture of rare genetic variants associated with relatively high disease risk as well as many common genetic variants associated with an individual small disease risk. In SCZ, these genetic variants are rare and include deletions of large chromosome regions, protein-truncating variants, small insertions, and deletions, as well as missense variants. Thus, SCZ is a highly polygenic disease, and the current understanding is that several genetic factors act together to contribute to development of SCZ in most cases. Therefore, unlike the monogenic effects seen in LQTS, calmodulin variants in SCZ likely act in concordance with numerous other genetic variants, potentially through several molecular pathways, where Ca_V_1.2 dysregulation is likely just one of them and only for a subset of variants.

There is a broad biological context to understanding a pathological role of calmodulin variants in the brain: Calmodulin is a key regulator of numerous ion channels (e.g. Ca_V_ and Na_V_ isoforms), receptors (e.g. NMDA and IP_3_ receptors), kinases (e.g. CaMKII), and phosphatases (e.g. CaN and PDE). As such, calmodulin modulates neuronal development, health, and synaptic firing (22). Studies from cell culture and mouse models show that depletion of specific *CALM* transcript variants prevents neuronal migration and brain development (46, 47), and competitive removal of calmodulin protein in developing neurons in *Drosophila melanogaster* disrupted axonal development in the neural growth cone (48). Furthermore, in a *Caenorhabditis elegans* model, we have demonstrated that human calmodulin variants from cardiac patients have neurological consequences (49, 50). As such, the impact of calmodulin variants in the brain may be multifaceted and highly complex and we have likely only seen the tip of the iceberg.

In conclusion, we identify calmodulin variants as a new risk factor in SCZ. Our mechanistic findings suggest that calmodulin variants from SCZ patients act through two different mechanistic paradigms. In one paradigm, SCZ variants impair calcium affinity and Ca_V_1.2 binding and regulation similarly to variants from LQTS patients, but with a much smaller effect size. In an alternative paradigm, SCZ variants enhance calcium affinity and Ca_V_1.2 binding but have no discernible functional impact on Ca_V_1.2. Therefore, we propose that both paradigms, but particularly the latter, act through other or additional molecular targets in the brain to those known from LQTS. Together, this study warrants an expansion of the phenotypic manifestation of calmodulin variants to include SCZ.

## Data Availability

All data produced in the present study are available upon reasonable request to the authors.

## Acknowledgements

We owe thanks to Giulia Monti for her help in setting up the RNAscope experiments. This work was supported by Lundbeck Foundation (R205-2017-134 to HHJ, R324-2019-1933 to MTO), Independent Research Fund Denmark (3031-00333A to MTO, 3160-00016A to MB), and Simon Fougner Hartmanns Family Foundation to MN.

